# Persistent health issues, adverse events of significant concern, and effectiveness of COVID-19 vaccination- findings from a real-world cohort study of healthcare workers in north India

**DOI:** 10.1101/2022.03.26.22272613

**Authors:** Upinder Kaur, Sapna Bala, Aditi Joshi, Noti Taruni Srija Reddy, Chetan, Mayank Chauhan, Nikitha Pedapanga, Shubham Kumar, Anurup Mukherjee, Vaibhav Mishra, Dolly Talda, Rohit Singh, Rohit Kumar Gupta, Ashish Kumar Yadav, Poonam Jyoti Rana, Jyoti Srivastava, Shobha Bhat K, Anup Singh, Naveen Kumar PG, Manoj Pandey, Kishor Patwardhan, Sangeeta Kansal, Sankha Shubhra Chakrabarti

## Abstract

**Background:** There is paucity of real-world data on COVID-19 vaccine effectiveness and safety from cohort designs. The current study aimed to evaluate vaccine performance during second wave in India. It also aimed to determine adverse events of significant concern (AESCs), and to ascertain the effect of vaccination on persistent health issues in individuals post COVID-19.

**Methods:** A cohort study was conducted from July-2021 to December-2021 in a tertiary hospital of north India. The primary outcome was vaccine-effectiveness against COVID-19. Secondary outcomes were AESCs, and persistent health issues in those receiving vaccine. Regression analyses were performed to determine risk factors.

**Results:** In 2760 healthcare workers (HCWs) included, 1033 COVID-19 events were reported. Around 6-17% vaccine effectiveness was observed against COVID-19 occurrence. One dose-recipients were at 1.6-times increased risk of COVID-19. Prior SARS-CoV-2 infection was a strong independent protective factor against COVID-19 (aOR 0.66). Full vaccination reduced moderate-severe COVID-19 by 57%. Those with lung disease were at 2.5-times increased risk of moderate-severe COVID-19. AESCs were observed in 1.3% including one case each of myocarditis and severe hypersensitivity. Individuals with hypothyroidism were at 5-times and those receiving vaccine after recovery from COVID-19 were at 3-times higher risk of persistent health issues.

**Conclusion:** COVID-19 vaccination reduced COVID-19 severity but offered marginal protection against occurrence. Relationship of asthma and hypothyroidism with COVID-19 outcomes necessitates focused research. Independent protection of prior SARS-CoV-2 infection was high and persistent health issues were common in individuals receiving vaccine post COVID-19. Recommendations of vaccinating those recovered from COVID-19 need further studies.

## 1. Introduction

COVID-19 vaccines have been a major deterrent against the raging pandemic. Concerted efforts by several global organizations have resulted in an unprecedentedly rapid development of several vaccine candidates which offer protection against SARS-CoV-2 infection and more notably against severe disease and death. However, the efficacy claims of widely used COVID-19 vaccines in the pivotal clinical trials have not been replicated fully in real world data. The pivotal trials of Pfizer-BioNTech, Moderna, Astra Zeneca (manufactured as COVISHIELD by Serum Institute of India), and Bharat Biotech (COVAXIN) were centred around the primary outcome of prevention of PCR positive SARS-CoV-2 infections.^1–4^ Post-authorization data for most of these leading vaccines has been predominantly derived from test negative case-control studies.^5–8^ The test negative design can control selection and information bias but does not effectively block the bias due to health seeking behaviour which differs between vaccinated and unvaccinated and is influenced by COVID-19 severity. Further, the appropriate use of this design requires baseline matching of the groups with respect to demographic characteristics, prior SARS-CoV-2 infection, comorbidities, and other variables.^9^ It is quite likely that the true estimates of vaccine effectiveness may be better calculated while studying a fixed population with similar exposure levels in a cohort design.^10^

Studies done so far have highlighted the effectiveness rates of vaccine at the time when immune protection against COVID-19 is expected to develop i.e 14 days after second dose and 21 days after first dose. Equally important to know however are the epidemiological attributes of COVID-19 and its patterns during early post vaccination period. Worth exploring is the effect of COVID-19 vaccination on persistent post-COVID health events, sometimes referred to as long COVID. This is particularly relevant in those with a history of natural SARS-CoV-2 infection prior to vaccination. To our knowledge the question of how post-recovery vaccination affects general health of these individuals has not been addressed. Moreover, though short-term safety analysis of COVID-19 vaccines in controlled settings have provided favourable results, the post approval period witnessed numerous case reports and series of serious adverse events and adverse events of special interest. These include reports of adverse cardiac events, thrombosis at atypical sites and new onset autoimmune diseases.^11–13^ The incidence and patterns of such adverse events need to be addressed in the real-world setting with equal emphasis to provide a better understanding of the benefit-risk ratio of COVID-19 vaccines and to stratify patients at risk of developing adverse events. This becomes more relevant owing to the high rates of breakthrough infections being reported from most regions. In this regard, a one year prospective observational safety study in vaccinated priority groups is being carried out by us since February 2021.

With this background, we conducted a cohort study to include the remaining healthcare workers in a major tertiary research and teaching hospital of north India with almost similar degree of exposures to the SARS-CoV-2 due to occupational reasons. The present study centres around two major objectives. It aimed to evaluate vaccine performance during the second wave in India, including in the immediate post-vaccination period. It also aimed to determine the occurrence of adverse events of significant concern in vaccinated individuals. Apart from providing the risk factors of COVID-19 occurrence and severity, for the first time, the study ascertains the real-world effect of vaccination on persistent health issues in individuals who had COVID-19 either prior to or following vaccination.

## 2. Materials and Methods

### 2.1. Study design and setting

This retrospective cohort study was conducted during the period of July 2021 to December 2021 in a tertiary hospital of North India. The second wave of the COVID-19 pandemic started from mid-March 2021, peaking around mid-April 2021 and reached the baseline by end of May 2021 (**Supplementary Figure 1**). The period from 16^th^ March 2021-31^st^ May 2021 was primarily selected for the estimation of real-world vaccine effectiveness against COVID-19.

### 2.2. Study participants

Newly recruited participants were health care workers working in the institute. Broadly, they included consultants (teaching faculty), resident doctors, nursing staff, paramedical staff, laboratory personnel and administrative staff. The participants belonged to modern medical, dental, Ayurvedic (Indian traditional medical), and nursing services. The HCWs available in the institute during working hours were contacted by the study team members as per planned survey schedule for entire institute and relevant medical data was collected in a pre-designed case report form. The HCWs who could not be contacted during three visits made on three different days by study team members due to any reason, or those who refused to participate were not included in the study. Also excluded from the study were those whose COVID-19 related, or COVID-19 vaccination related details were incomplete. The participants were identified as ‘confirm’ or ‘suspect’ COVID-19 cases as per Ministry of Health and Family Welfare guidelines (together labelled as COVID-19 cases).^14^ These cases were compared to those with no COVID-19 like events for determination of risk factors of incidence. Severity of COVID-19 was also decided as per MoHFW guidelines.^14^ Participants with moderate-severe forms of COVID-19 were clubbed in one group and analysed in determining risk factors of COVID-19 severity. HCWs who were symptomatic during the study period but negative in RT-PCR lab reports were excluded from the analysis. To avoid survival bias, information was collected from each department regarding any deaths of employees during the study period, and family members were contacted to extract the details of vaccination status and mortality of these individuals.

### 2.3. Ethical permission

The study started after obtaining ethical permission from the Institute Ethics Committee. Written informed consent was obtained from all the study participants.

### 2.4. Data sources/ measurement

All medical data was collected in a pre-designed case report form. The data pertaining to relevant demographics, medical history, concomitant drug history, any history of SARS-CoV-2 infection in the past, COVID-19 vaccination history, adverse events following immunization (AEFIs) and COVID-19 related medical details were collected. For those vaccinated, participants were recruited irrespective of the type of vaccine and centre at which vaccine was received. Case report forms which lacked clarity on COVID-19 or vaccination status were excluded.

### 2.5. Outcomes measured

#### 2.5.1. Primary outcomes

The primary outcome of the present study was real-world vaccine effectiveness against COVID-19 occurrence and severity during the second wave. We also aimed to predict determinants of both occurrence and severity of disease.

To generate more robust data on real-world performance of the vaccines, we used multiple analytic designs, in a departure from previously reported literature. Evaluation was performed at various time points as detailed below.

Three strategies were adopted for analysis of vaccine effectiveness-

##### Strategy A (Definition A)

The standard definition of fully vaccinated and partially vaccinated as used in the pivotal clinical trials was used as definition A. Individuals were categorized as ‘2’ dose recipients if they had received their second dose before >14 days of reference date, and as ‘1’ dose recipient if they had received their first dose before >21 days of reference date. Reference date of calculating these time intervals for those developing COVID-19 was either the date of laboratory diagnosis of COVID-19 or the date of onset of symptoms, whichever was earlier. The reference date of calculating time intervals for those not developing COVID-19, was fixed on 12^th^ April 2021 (peak of second wave as per **Supplementary Figure 1**). This strategy was employed as, in case onset of second wave or end of second wave had been taken as reference date instead of 12^th^ April, there could have been potential underestimation or overestimation of vaccine effectiveness, which we wanted to avoid. Individuals were labelled as recipients of ‘0’ dose if no vaccine dose was received before occurrence of COVID-19 or the date of 12^th^ April 2021.

##### Strategy B (Definition B)

This strategy was considered because there have been some reports of transient immunosuppression after receiving the first dose of the vaccine. To correct for this period, in definition B, participants were labelled as ‘2’ dose recipients if the second dose had been received before >14 days of the reference date, and were labelled as ‘1’ dose recipients if first dose had been received at any time before the reference date. As in definition A, reference dates for calculating these time intervals for those developing COVID-19 was either the date of laboratory diagnosis or the date of onset of symptoms, whichever was earlier. The reference date of calculating time intervals for those not developing COVID-19, was fixed on 12^th^ April 2021 (**Supplementary Figure 1**). The ‘0’ dose group included those who did not receive any vaccine dose before occurrence of COVID-19 or the set date of 12^th^ April 2021.

##### Strategy C: Pure comparison between unvaccinated HCWs and those fully vaccinated before the start of second wave

This strategy was adopted to give vaccine effectiveness estimates which would be unadulterated by any vaccinations received during the second wave, any hypothetical transient immunosuppression, and any accidental super-spreader events from vaccination centres.

For this comparison, we considered as one group those HCWs who were fully vaccinated before the start of the second wave and as comparator group, those who were unvaccinated till the wave touched the baseline (31^st^ May 2021). All participants who had received any dose of vaccine during the period of study (period of second wave from 16^th^ March 2021 to 31^st^ May 2021) were excluded from this analysis. For this analysis, since the start of second wave was selected as 16^th^ March 2021, to be considered fully vaccinated, participants must have received their second dose by 1^st^ March 2021.

#### 2.5.2. Secondary outcomes

Adverse events of significant concern (AESCs) was the main secondary outcome. Events were graded using the FDA adverse event severity grading scale and categorized under AESC if there was occurrence of any of:

1. Any Serious AEFI
2. Any Severe AEFI (FDA grade 3)
3. Any moderate-severe AEFI (FDA grade 2-3) which persisted for ≥7 days
4. Any moderate AEFI (FDA grade 2) which persisted for ≥ 4 weeks
5. Any mild-moderate AEFI (FDA grade 1-2) that persisted for ≥ 12 weeks

Causality association was performed for all serious AEFIs using the World Health Organization (WHO) scale of causality assessment.

Other secondary outcomes included health issues in the participants at the time of visit made by study team. These events were studied individually and those persisting for at least 2 months were explored for any association with history of COVID-19 or COVID-19 vaccine. MedDRA classification was used to assign System Organ Class (SOC) to these events. Broadly, the events were categorized into four groups:

A. Vaccine (post COVID-19) associated: If health events were reported in those who received COVID-19 vaccine after recovering from natural SARS-CoV-2 infection in the past.
B. COVID-19 (post vaccine) associated: If events were reported in those who developed COVID-19 after receiving COVID-19 vaccine.
C. COVID-19 associated: If events were reported in unvaccinated individuals who developed COVID-19.
D. Vaccine associated: If events were reported in vaccinated individuals with no history of COVID-19 till date of enrollment.

### 2.6. Sample size

The sample size estimation was based on the major primary outcome of the study i.e. vaccine effectiveness. Considering 0.6% rate of occurrence of any symptomatic COVID-19 in vaccinated and 1.9% rate of occurrence of any symptomatic COVID-19 in the control group (based on Voysey et al)^3^, α of 0.05 and power of 80%, the minimum sample size required for the present study was 2286. Considering a 5% rate of exclusion of participants because of incomplete information, a minimum sample size of 2400 was required. The data collection was stopped on 31^st^ December 2021 by which time 2765 participants had been enrolled.

### 2.7. Statistical analysis

Data was represented as frequencies for dichotomous variables and as mean and median values for continuous variables, depending upon skewness. Frequencies and percentages are provided for participants developing AESCs. Chi square test was applied to assess association between dichotomous variables and occurrence as well as severity of COVID-19 and also with post-COVID-19 persistent symptoms. Time to occurrence of COVID-19 between vaccinated and unvaccinated groups was compared using the Kaplan-Meier survival analysis. This was followed by Cox proportional hazard model to predict the risk factors of COVID-19 occurrence after adjusting for potential covariates. To determine risk factors of severity of COVID-19 and risk factors of post-COVID persistent symptoms, binary logistic regression analysis was used. The variables with P < 0.05 in unadjusted bivariate analysis were selected for Cox-proportional and logistic regression models.

## 3. Results

### 3.1. Descriptive Data

Overall, medical data was collected successfully from 2765 HCWs. After excluding case report forms of 5 HCWs which lacked essential vaccine related or COVID-19 related information, a total of 2760 HCWs were enrolled in the study. **Figure 1** shows flowchart of selection of participants and steps followed in each analysis. Mean age of HCWs was 34.9 (±9.9) years (male=1740, female=1020). After excluding 69 HCWs who were RT-PCR negative suspects, 2691 HCWs were included for main analysis. 1033 COVID-19 events were identified in them from the period of February to December 2021. Six HCWs reported developing COVID-19 twice in this period. A total of 973 events occurred in 969 HCWs in the second wave or analysis period (16^th^ March to 31^st^ May 2021). Of these 973 events, 238 were rated as moderate-severe grade.

**Figure 1.**
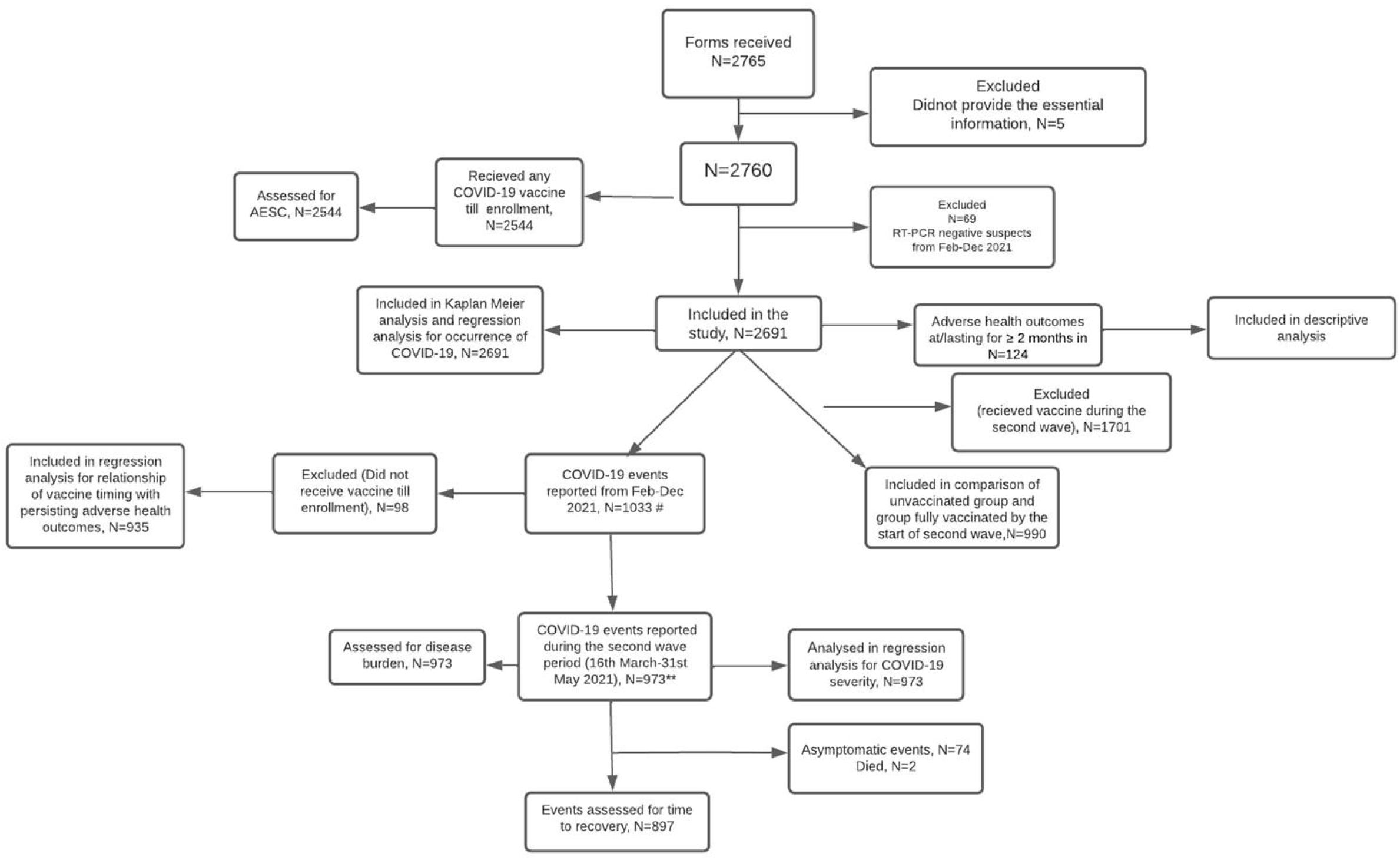
STROBE Flow diagram of selection of participants and steps followed for each analysis.

### 3.2. Main Results

#### 3.2.1. Occurrence of COVID-19

In unadjusted analysis, age, sex, prior history of COVID-19, presence of hypothyroidism and vaccination status shared a statistically significant association with occurrence of COVID-19 and were selected for Cox-proportional hazard analysis. Occurrence of COVID-19 was common in HCWs who were young, females and had a history of hypothyroidism. No significant difference in COVID-19 occurrence was seen between vaccinated and unvaccinated groups when assessed as per standard definition of vaccination status i.e., strategy A. Interestingly, COVID-19 occurred more commonly in vaccinated individuals compared to unvaccinated when assessed as per strategy B which considered the probabilities of vaccination centers being super-spreader sites or a potential transient immune suppression following the vaccination. The occurrence of COVID-19 was less common in HCWs with prior history of SARS-CoV-2 infection (p<0.001).

Figure 2. shows comparison of time to occurrence of event (COVID-19) between vaccinated and unvaccinated groups. Individuals belonging to ‘1’ dose group developed COVID-19 earlier compared to the unvaccinated and ‘2’ dose group with statistical significance. After adjusting for potential confounders in Cox proportional hazard model, age, sex, prior history of COVID-19 and vaccination status emerged as tentative determinants of occurrence of COVID-19 (**Table 1a**). The risk of occurrence of COVID-19 was nearly 1.5-times for those <40 years of age as compared to participants ≥ 40 years, 1.2-times higher for females compared to males and 1.6-times higher in ‘1’ dose group compared to the unvaccinated. Prior SARS-CoV-2 infection was an independent protective factor with a 34% lower risk of COVID-19 in this group compared to those with no prior history of SARS-CoV-2 (p<0.001).

**Table 1:** Regression analyses to determine tentative risk factors for occurrence and severity of COVID-19. **[2a: Cox-proportional hazard model for risk factors of occurrence; 2b: Binary logistic regression analysis for risk factors of moderate-severe COVID-19** aHR: adjusted hazard ratio; aOR: adjusted odds ratio **\***As per definition B (P<0.05 in unadjusted bivariate analysis, only for definition B) **As per definition A. With both definitions of vaccination status showing P < 0.05 in unadjusted bivariate analysis, the standard definition (definition A) was chosen for logistic regression analysis. Similar statistical results were seen even with definition B]

| 1a |  |  | 1b |  |  |
| --- | --- | --- | --- | --- | --- |
| Tentative risk factors (n=2691) | aHR | P-value | Tentative risk factors (n=973 COVID-19 events) * | aOR | P-value |
| **Age**<br><40<br>≥40 (Reference) | **1.46** (1.25-1.72) | **<0.001** | **Pre-existing lung disease**<br>Yes<br>No (Reference) | **2.54** (1.13-5.7) | **0.02** |
| **Sex**<br>Female<br>Male (Reference) | **1.23** (1.08-1.4) | **<0.002** |  |  |  |
| **Prior history of COVID-19**<br>Yes<br>No (Reference) | **0.66** (0.54-.081) | **<0.001** | **Vaccination status\*\***<br>2 doses<br>1 dose<br>0 dose (Reference) | **0.43** (0.31-0.60)<br>0.83 (0.54-1.26) | **<0.001**<br>0.38 |
| **Hypothyroidism**<br>Yes<br>No (Reference) | 1.31 (0.97-1.77) | 0.07 |  |  |  |
| **Vaccination status\***<br>2 doses<br>1 dose<br>0 dose (Reference) | **1.17** (1.01-1.37)<br>**1.58** (1.33-1.9) | **0.04**<br>**<0.001** |  |  |  |

#### 3.2.2. Severity of COVID-19

Pre-existing lung disease and vaccination status were found to be associated with moderate-severe forms of COVID-19 with statistical significance. Disease burden as well as time to recovery from COVID-19 were significantly lower in ‘2’ and ‘1’ dose groups compared to ‘0’ dose group. Median time to recovery was 10 (7,18) days in ‘0’ dose group and 7 (5,14) days for ‘2’ dose group (p=0.002). Disease burden expressed as median number of symptoms was 5 (3,7) in ‘0’ dose group and 3 (2,5) in ‘2’ dose group (p<0.001).

**Figure 2.**
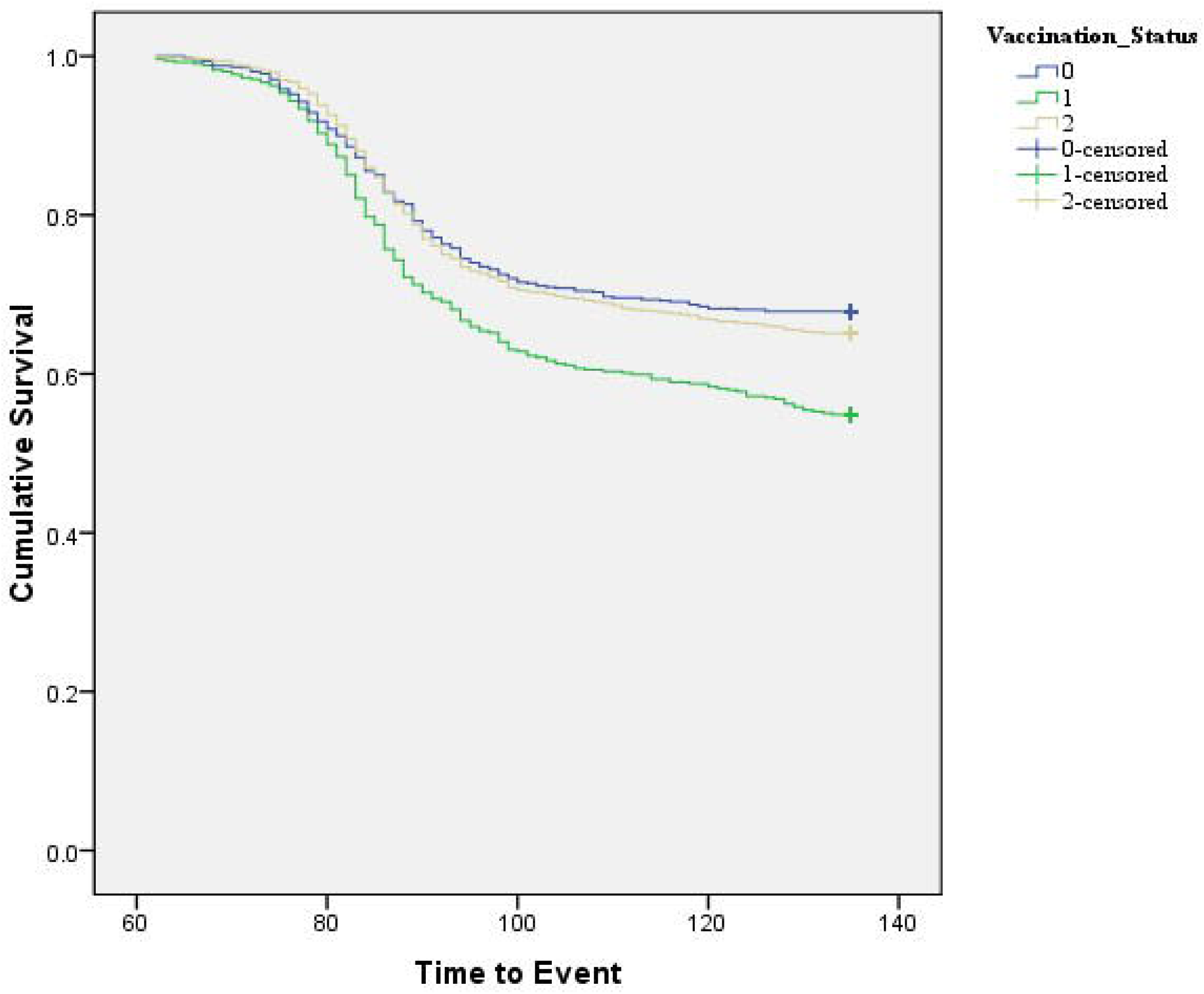
Kaplan Meier curve showing time to occurrence of event (COVID-19) in “1” dose and “2” dose vaccinated and unvaccinated groups.

Binary logistic regression analysis showed preexisting lung disease to be associated with 2.5-times higher odds of moderate-severe COVID-19 (**Table 1b**). Compared to the unvaccinated, the HCWs in ‘2’ dose group were at 57% lower risk of suffering from moderate-severe COVID-19.

#### 3.2.3. Persistent health issues

A total of 935 COVID-19 events (between February to December 2021) were assessed for relationship between timing of COVID-19 vaccination and health issues persistent for ≥ 2 months. With statistical significance, persistent health issues were common in ≥40 years age group. Vaccinated HCWs with a history of hypothyroidism, inflammatory arthritis, diabetes mellitus or allergy were more likely to have persistent health issues. Persistent health issues were also common in recipients of COVAXIN compared to those receiving COVISHIELD (p=0.03). Interestingly, a higher percentage of HCWs who received the vaccine after natural COVID-19 had persistent health issues compared to those who received the vaccine before COVID-19. **Table 2** shows results of logistic regression and validates potential risk factors of persistent health issues in HCWs. After adjusting for potential confounders, presence of hypothyroidism was associated with 5-times higher risk of persistent health issues and history of allergy was associated with 2-times higher risk. Receiving the vaccine after natural SARS-CoV-2 infection of second wave was associated with a nearly 3-times higher risk of persistent health events.

**Table 2.** Regression analysis to determine risk factors for persisting health issues in health care workers with history of COVID-19 vaccination before or after COVID-19 between February to December 2021.

| **Tentative risk factor** | **Odds ratio** | **P-value** |
| --- | --- | --- |
| **Vaccination after COVID-19**<br>Yes<br>No (reference) | **2.8** (1.8-4.4) | **<0.001** |
| **Age (years)**<br><40<br>≥40 (reference) | 0.67 (0.39-1.15) | 0.15 |
| **Diabetes mellitus**<br>Yes<br>No (reference) | 1.1 (0.44-2.7) | 0.85 |
| **Autoimmune arthritis**<br>Yes<br>No (reference) | 30 (3-303) | 0.004 |
| **History of allergy**<br>Yes<br>No (reference) | **2** (1.2-3.4) | **0.01** |
| **Hypothyroidism**<br>Yes<br>No (reference) | **5.25** (2.6-10.5) | **<0.001** |
| **Vaccine type**<br>COVAXIN<br>COVISHIELD | 0.97 (0.2-4.4) | 0.97 |

##### 3.2.3.1. Descriptive data of persistent health issues

A total of 124 HCWs reported health events which were persisting till date of interview and for at least two months. The MedDRA classification of such events along with their association with COVID-19 or COVID-19 vaccine is shown in **Figure 3**. Majority (n=74, 59%) of the HCWs reporting persistent health events had received the vaccine after natural SARS-CoV-2 infection and belonged to the ‘vaccine (post COVID-19)’ group. When classified as per MedDRA, majority of the persistent health issues (in 124 HCWs) belonged to the SOC of ‘general disorders and administration site conditions’ (n=35) followed by ‘musculoskeletal and connective tissue disorders’ (n= 22) and ‘cardiac disorders’ (n=17).

**Figure 3.**
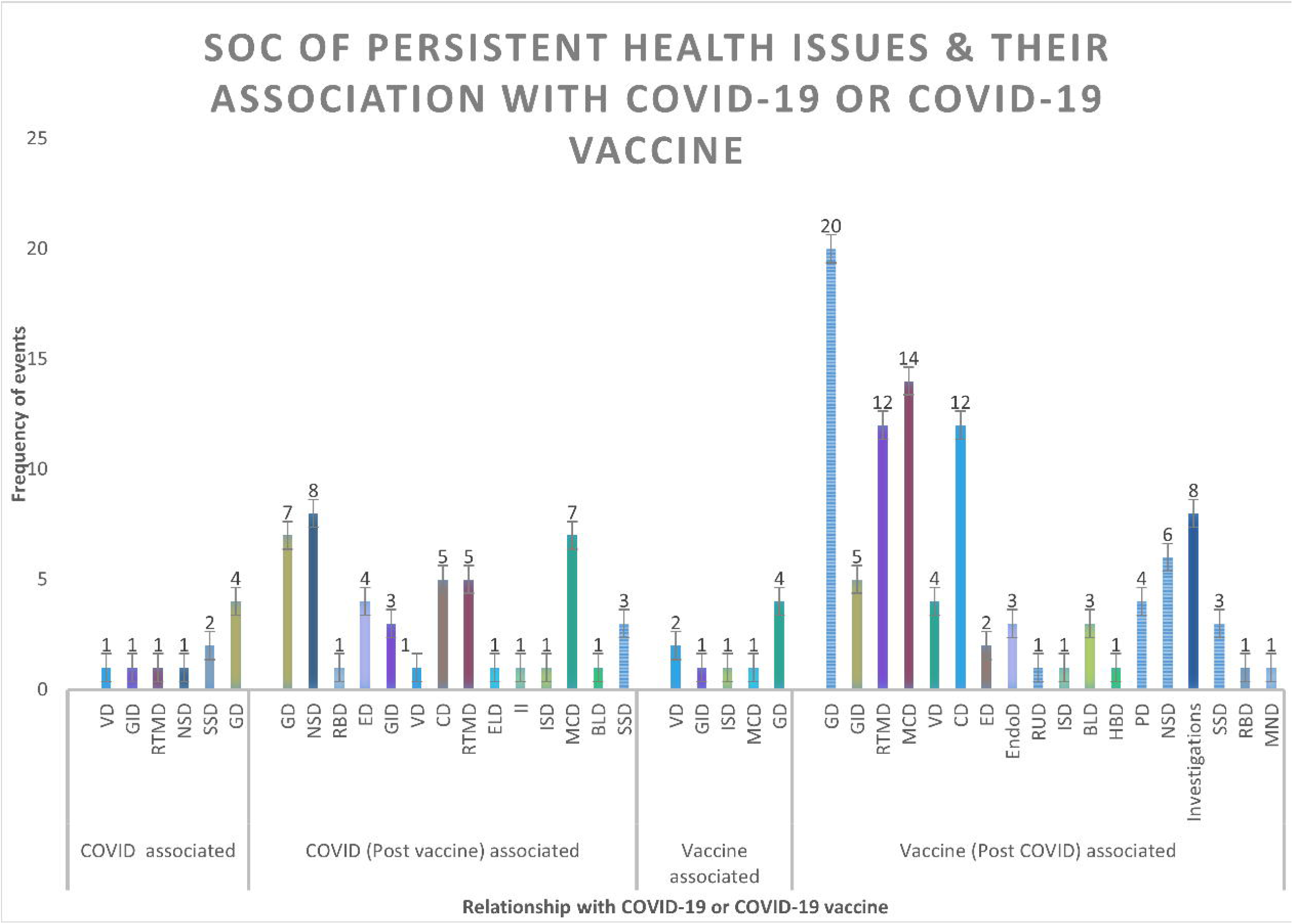
System organ class (SOC) of persistent health issues and their relationship with COVID-19 or COVID-19 vaccine.

#### 3.2.4. Adverse events of significant concern (AESCs)

Out of the total 2544 HCWs receiving any vaccine at any time till date, 33 HCWs developed AESCs (1.3%). A total of seven HCWs developed AEFIs of ‘serious’ grade (0.3%). Eight HCWs developed ‘severe’ AEFIs, six had AEFIs of moderate-severe grade that persisted for ≥1 week, eight had ‘moderate’ AEFIs that persisted for ≥4 weeks and four had mild-moderate AEFIs which lasted for ≥12 weeks. These AESCs persisted with no or partial recovery in 13 HCWs. One HCW had a miscarriage and one died due to a cardiac event. Two HCWs recovered while on new treatment and full recovery was seen in the remaining 16 HCWs. Time to recovery varied from 3-150 days. Of the three deaths reported in our study, one death occurred due to cardiac arrest in a male patient in his 40s with underlying obesity, uncontrolled hypertension, and diabetes. The deceased had received his first dose of vaccine around 1.5 months before the event. The remaining two deaths occurred in unvaccinated individuals, one with underlying hypertension, and the other with underlying diabetes (one confirm COVID-19, one suspect COVID-19).

## 4. Discussion

The design and conduct of the present study were preceded and influenced by a published preliminary study of ours showing high rates (27-46%) of occurrence of COVID-19 in vaccinated priority groups. Nearly 40% breakthrough infection rates were reported by the authors in doctors, majority of whom are currently employed in the institute where the present study was planned. To make comparisons with the HCWs who were unvaccinated at the time of the pandemic and to generate region specific vaccine effectiveness, it was decided to include remaining HCWs of the institute to get a larger vaccinated and unvaccinated cohort.

Vaccine effectiveness rates of 60-80% have been shown in clinical trials, and in majority of the published real-world studies on ChAdOx1-nCoV-19 and inactivated SARS-CoV-2 vaccines.^3–6^ On the contrary, a marginal COVID-19 protection rate (<6%) was observed with COVID-19 vaccines in the present study even after following the standard definitions of timings of immune protection which have been used in all pivotal trials of COVID-19 vaccines. These wide variations can be explained to some extent by the study designs employed in majority of the post approval vaccine studies. The test negative case control designs on which the published studies were based categorize the individuals to cases and controls depending upon the results of laboratory tests. These designs effectively control the selection bias of symptomatology based traditional case-control studies. Unchecked however are the differences in health seeking behavior due to variable presentations and severity of disease in individuals. The design should not be used in situations where patterns of presentation of disease vary in cases and controls. Baseline characteristics of cases and controls also need to be matched in the absence of which potential errors are generated in the estimation of vaccine effectiveness rates. Further, since test negative designs rely only on laboratory tests, the sensitivity and specificity of the laboratory assay becomes a major determining factor in case identification. Many of the post-approval studies have had these limitations.

The maximum vaccine protection observed after excluding the participants who received any dose during the second wave (pure vaccinated vs unvaccinated cohort analysis performed in Strategy C), was close to 17%. Interestingly, ‘1’ dose recipients showed a higher rate of SARS-CoV-2 infection compared to the unvaccinated. The time to occurrence of COVID-19 in “1” dose recipients was significantly shorter, as corroborated from the findings of the Kaplan-Meier analysis. The high risk of acquisition of disease in ‘1’ dose vaccinated individuals persisted even after adjusting for potential confounders in the Cox proportional hazard model. Compared to the unvaccinated, “1” dose recipients were nearly at 1.6-times higher risk of developing COVID-19. Increased COVID-19 occurrence rate after the first dose has been observed in some other studies and has been linked to the underlying high-risk group of the participants enrolled as well as to the vaccination centers being super-spreading sites of infection.^5,6^ However, since majority of the observed COVID-19 infections in our study were breakthrough, a direct immunomodulatory action of COVID-19 vaccines should also be investigated as a potential cause of suboptimal vaccine protection and increased propensity towards COVID-19. Some evidence in this regard is provided by a detailed Chinese study on post-vaccination immune modulation.^15^

Among other factors, young individuals <40 years of age, and females were observed to be at higher risk of acquiring COVID-19 with respect to comparators. Since majority of the participants in the present study were young health care workers, the age specific findings cannot be extrapolated to the general population. The regression analysis showed prior history of COVID-19 as a strong independent protective factor associated with lower rates of disease. Nearly 34% lower risk of COVID-19 was observed in individuals with SARS-CoV-2 infection in the past.

With respect to severity of COVID-19, vaccine effectiveness of 46-51% and 13-19% were observed with ‘2’ and ‘1’ dose respectively. These rates are lower than the severity benefits claimed in controlled settings and some real-world studies.^3,16^ However, the protection offered by vaccines remained statistically significant in regression analysis after adjusting for potential confounders. Compared to unvaccinated, fully vaccinated individuals were at 57-60% lower risk of moderate-severe disease. This is similar to reports by another group from North India.^17^

Apart from vaccination status, presence of pre-existing lung disease, particularly asthma was associated with 2.5-times higher odds of moderate-severe COVID-19. Majority of participants with asthma enrolled in this study had disease of mild-moderate severity and were controlled either on inhaled corticosteroids or systemic leukotriene antagonists. The evidence associating asthma to COVID-19 is conflicting at present. Contrary to expected, asthma was observed to be an underrepresented co-morbidity in hospitalized COVID-19 individuals.^18^ In a large cohort study using electronic health records of patients in England, mild-moderate asthma not requiring systemic steroids was not associated with worse clinical outcomes.^19^ Only a modest risk of poor COVID-19 outcomes (aHR 1.13) existed with severe asthma. It has been suggested that the Th phenotype of patients should be explored to delineate the relationship of asthma with COVID-19.^20^

Another important objective of the present study was to predict the determinants of post-COVID-19 persistent health issues, and to shed light on the safety profile of COVID-19 vaccines after a natural SARS-CoV-2 infection. After adjusting for potential confounders, presence of hypothyroidism was associated with more than 5-times higher risk of post-COVID persistent health issues, compared to those with euthyroid state. A nearly 2-times higher risk was evident in individuals with a history of allergy to any stimuli. Interestingly, a close to 3-times higher risk of post-COVID persistent health issues was observed in individuals receiving the vaccine after past natural SARS-CoV-2 infection in 2021, compared to those who received the vaccine before getting the infection. Presence of inflammatory arthritis was also associated with higher risk of post-COVID persistent symptoms, though the confidence intervals varied widely and the overall number of individuals with immune mediated arthritis was small. The high risk of persistent health issues seen with COVAXIN in unadjusted bivariate analysis did not remain statistically significant in regression analysis. The associations of all these proposed risk factors were corroborated in a separate regression analysis extended to involve any SARS-CoV-2 infection in the past. The increased risk of persistent health issues in those vaccinated post-recovery from SARS-CoV-2 has not been explored in the real world. Most studies have focused on antibody titer augmentation and have suggested a potential booster effect. However, this needs to be viewed realistically considering the additive burden of adverse events or persistent health issues. In light of much new evidence that vaccine induced immunity is short lasting, vaccinating those recovered from natural infection needs close scrutiny.

A detailed safety analysis was performed in individuals with health issues persistent for ≥2 months. Majority (60%) of them had received the vaccine post-recovery from natural SARS-CoV-2 infection. Nearly 27% of persistent health issues were related to developing COVID-19 post vaccination. Adverse events of significant concern (AESCs) developed in 33 participants, giving the AESC rate close to 1.3%. Serious AEFIs occurred in 0.3% of HCWs. Considering AESCs with ‘probable’ causality association with the vaccine, the incidence of cardiac events following COVID-19 vaccines and that of severe hypersensitivity reactions comes close to 1 per 2544 cases which is much higher than what has been claimed by vaccine manufacturers. Of the three deaths reported, one occurred due to cardiac arrest in a partially vaccinated individual with multiple co-morbidities and the remaining two deaths occurred in unvaccinated comorbid individuals, possibly due to COVID-19.

### 4.1. Limitations

The possibility of recall bias exists in the study due to its retrospective design. However, the participants being healthcare workers, the information may be considered mostly reliable. As the study is based on a predominantly healthy younger population, the results may not be extrapolated to the general Indian population. Because of the small number of death events, and no autopsy details available in this questionnaire-based study, not much interpretation can be made in regard to this either. The study further does not provide virus variant specific information.

## 5. Conclusion

COVID-19 vaccination provided a marginal protection against the occurrence of COVID-19 and a modest protection against the severity of disease. Compared to the unvaccinated, a high risk of occurrence of COVID-19 was observed in participants receiving ‘1’ dose of vaccine. Previous infection by SARS-CoV-2 acted as an independent protective factor against COVID-19 occurrence. Preexisting lung disease, mainly asthma independently enhanced the risk of moderate-severe COVID-19 and necessitates a focused study of COVID-19 vaccines in individuals with asthma. Receiving any dose of vaccine after natural SARS-CoV-2 infection was associated with higher risk of persistent health issues.

With considerably lower protection against COVID-19 than predicted from controlled settings and more than double rates of chronic adverse events in those vaccinated after a natural history of SARS-CoV-2 infection, the authors suggest that the concept of vaccinating recovered individuals may need closer scrutiny. It may not be beneficial in the light of multiple breakthrough infections, and short-lasting immunity post-vaccination.

Vigilance for prolonged health events is warranted in individuals with hypothyroidism and history of allergy, and those receiving any COVID-19 vaccine after natural SARS-CoV-2 infection. Patients of inflammatory arthritis also need to be monitored for long term health events post COVID-19 vaccine. The incidence of severe hypersensitivity reactions and myocarditis seems to be higher than what has been claimed, and warrant studies focussed on long term vigilance of vaccines to safeguard public health.

## Data Availability

All data produced in the present study are available upon reasonable request to the authors

## Acknowledgements

None

## Ethical Statement

No human experimentation performed. All procedures performed as per the Declaration of Helsinki and its further modifications. Written informed consent obtained from all participants. Study conducted after permission from Institute Ethics Committee of the Institute of Medical Sciences, Banaras Hindu University.

## Funding Statement

No funding support.

## Conflicts of Interest

None of the authors report any conflict of interest.

## Figure Legend

**Supplementary Figure 1. Date-wise COVID-19 case counts during second wave in study setting**

## Notes

### Competing Interest Statement

The authors have declared no competing interest.

### Funding Statement

This study did not receive any funding

### Author Declarations

The Institute Ethics Committee of the Institute of Medical Sciences, Banaras Hindu University, Varanasi, India

