## Supplementary figures and images for "Persistent health issues, adverse events of significant concern, and effectiveness of COVID-19 vaccination- findings from a real-world cohort study of healthcare workers in north India"

### Supplementary Figure 1

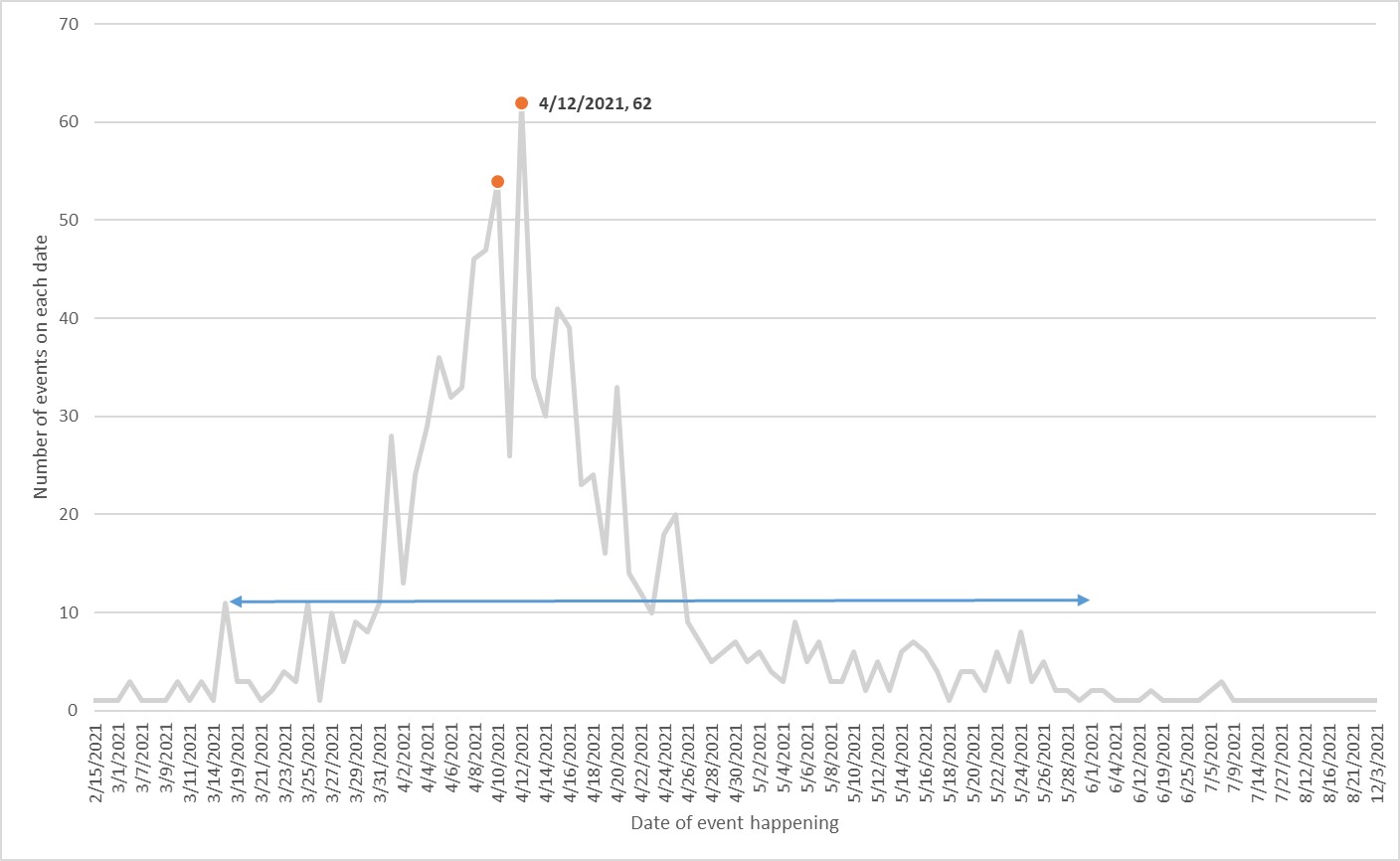
